# Causes And MEchanisms foR non-atopic Asthma in Children (CAMERA) Study: rationale and protocol

**DOI:** 10.1101/2025.01.20.25320833

**Authors:** Mary Njoroge, Gabriela Pimentel Pinheiro, Cinthia Vila Nova Santana, Hajar Ali, Stephanie Hobbs, Santiago Mena-Bucheli, Natalia Romero-Sandoval, Steven Robertson, Charlotte E Rutter, Donna Davoren, Collin Brooks, Jeroen Douwes, Philip J Cooper, Harriet Mpairwe, Camila A Figueiredo, Alvaro A Cruz, Mauricio L Barreto, Neil Pearce, Lucy Pembrey, the CAMERA study group

**Affiliations:** Department of Medical Statistics, London School of Hygiene and Tropical Medicine; MRC/UVRI and LSHTM Uganda Research Unit, Entebbe, Uganda; Institute of Health Sciences, Federal University of Bahia, Salvador, Brazil; Centre for Public Health Research, Massey University, Wellington, New Zealand; Fundacion Ecuatoriana Para Investigacion en Salud, Quito, Ecuador; School of Medicine, Universidad Internacional del Ecuador, Quito, Ecuador; Institute of Infection and Immunity, St George’s University of London, London, UK; ProAR Foundation and Federal University of Bahia, Salvador, Brazil; The Centre for Data and Knowledge Integration for Health (CIDACS), Fiocruz, Bahia, Brazil

## Abstract

The Causes And MEchanisms foR non-atopic Asthma in children (CAMERA) study was designed to investigate risk factors and mechanisms of non-atopic asthma in children and young adults in Brazil, Ecuador, Uganda, and New Zealand. Initial epidemiological analyses using existing datasets identified and compared risk factors for both atopic and non-atopic asthma. The focus of this paper is the protocol for sample collection and analysis of clinical data on possible non-atopic mechanisms.

In each of the four centres, the CAMERA study will enroll 160 participants aged 10 – 28 years, equally distributed among atopic asthmatics (AA), non-atopic asthmatics (NAA), atopic non-asthmatics and non-atopic non-asthmatics. Participants will be new recruits or returning World ASthma Phenotypes (WASP) study participants.

Phase I consists of skin prick tests to define atopy, a general CAMERA questionnaire that covers respiratory and general health to identify asthma cases, followed by an asthma control questionnaire for asthmatics only. Phase II consists of a stress questionnaire and the following clinical assessments: lung function, nasal cytology, blood sampling, *in vitro* whole blood stimulation to assess IFN-γ production, hair cortisol concentration, dry air and capsaicin challenges, plus in a subset, cold air challenges. Analyses will compare inflammatory, physiological and clinical parameters across the four groups overall and by country.

Here, we present the protocol for the CAMERA study, to provide relevant methodological details for CAMERA publications and to allow other centres globally to conduct similar analyses. The findings of this mechanistic multi-centre study will inform new and phenotype-specific prevention and treatment approaches.

## Introduction

Childhood asthma is a major global public health burden with increasing prevalence in low-and-middle-income-countries (LMICs) (1), and generally high (but varied) asthma prevalence across high-income countries (HICs) (2). Asthma is now considered to be a heterogenous disease associated with multiple causal pathways that result in similar clinical effects including respiratory symptoms and variable airways obstruction (3). The mechanisms underlying different asthma phenotypes vary, and those associated with non-atopic disease remain poorly understood.

The concepts of atopic and non-atopic asthma have evolved since their introduction in the late 1940s by Rackemann, who referred to them as extrinsic and intrinsic asthma respectively (4). Atopic asthma (AA) is classified based on symptoms triggered by common allergens, and confirmed by serum specific IgE or a positive skin prick test (SPT) to relevant aeroallergens. It is often associated with Th2-mediated eosinophilic airway inflammation (5,6). On the other hand, the mechanisms underlying non-atopic asthma (NAA) (7), remain unclear. While studies in the 1990s demonstrated elevated Th2 cytokines in some cases, non-Th2 pathways are also implicated, including neutrophilic inflammation associated with Th1 and Th17 responses and increased levels of IL-8, IL-17A, and IL-1β in certain patient subgroups (8). However, there is often no clear evidence of any airway inflammation (i.e. paucigranulocytic), and, paradoxically, eosinophilic airway inflammation may be present (9). NAA can be exacerbated by triggers such as exercise and environmental exposures such as cold and dry air rather than allergens (10). Even though neural pathways, non-atopic, non-Th2 inflammatory processes and chronic stress are possible mechanisms underlying NAA, evidence for this is scant. This is likely to be because NAA has been relatively understudied; possibly due to high AA prevalence in HICs and the prevailing dogma over several decades that asthma *is* an allergic condition. However NAA is relatively common in LMICs compared to HICs (11). It is therefore important to understand the causes and mechanisms of NAA to identify potential risk factors and elucidate underlying mechanisms to enable the development of targeted disease management approaches in both HICs and resource-limited LMIC settings.

Our previous World Asthma Phenotypes (WASP) study found that asthma characteristics in LMICs and HICs varied substantially; around half were non-eosinophilic in New Zealand (NZ) and United Kingdom and around two-thirds were non-eosinophilic in Ecuador, Brazil and Uganda (12). Initial work on the CAMERA study using existing datasets showed that although atopy increases asthma risk, risk factors for developing asthma were similar in atopic and non-atopic people. However, as the mechanisms of NAA are not well understood, there could be other important risk factors that were not captured (13). Studies have increasingly focused on understanding the underlying mechanisms of NAA in addition to identifying risk factors. For example, previously reported data from Brazil showed that interferon-gamma (IFN-γ) production in House dust mite (Dermatophagoides pteronyssinus) allergen-stimulated peripheral blood leukocytes was higher in NAA than in AA (14). Given that non-eosinophilic asthma (based on sputum) and NAA overlap by approximately 60-70% (15,16), and Chen et al. reported overlap in both inflammatory and clinical characteristics (17), we will analyse inflammatory phenotypes (using nasal lavage) and atopic status (using SPT) to better characterise immunopathology in these distinct but related phenotypes. To investigate non-inflammatory mechanisms, we will use specific challenge tests: dry/cold air challenges to assess airway hyperresponsiveness, and capsaicin challenges to evaluate sensory nerve reactivity. A small study by Ali et al. reported enhanced capsaicin sensitivity among non-eosinophilic asthmatics compared to non-asthmatics, with a similar trend observed when compared to eosinophilic asthmatics (but only weak evidence of a difference for the latter). Notably, capsaicin sensitivity was not associated with atopic status, highlighting the potential role of sensory nerve reactivity in the pathophysiology of NAA (17). The CAMERA study will clarify the role of these potential mechanisms by assessing a wider range of participants from varying settings with respect to geography, asthma prevalence, socioeconomic status, ethnicity, and environmental exposures.

The CAMERA study therefore builds on the WASP study findings and framework to investigate possible neural and non-neural mechanisms relevant to NAA across diverse populations (Figure S1).

Here, we present the rationale, design, and study procedures for the CAMERA study clinical investigations.

### Study Protocol

#### Study centres, groups and recruitment

CAMERA is a cross-sectional design that will enrol children, adolescents and young adults aged 10 – 28 years; recruitment methods and ages vary by centre. Brazil (12 – 28 years) will recruit from the previous WASP study and the Programme for Control of Asthma in Salvador (ProAR) database. Ecuador (10 – 24 years) will recruit new participants with asthma from a public specialist hospital registry in Portoviejo, Manabi province, and in the community where patients live (1 per case). Uganda (13 – 23 years) will recruit WASP participants and additional participants from primary, secondary, and tertiary schools in Wakiso district, Entebbe. New Zealand (16 – 22 years) will recruit from the WASP study population and local community (schools, universities and medical centres).

Each centre will enrol 160 eligible participants to obtain four groups of 40 participants representing AA, NAA, atopic non-asthmatics, and non-atopic non-asthmatics. Atopy will be defined by SPT positivity to a standardised allergen panel (Table S1) using WASP protocol methodology (18).

#### Identification of asthma cases and non-asthmatics

Asthma cases will be identified on the basis of current asthma symptoms and/or medication-use in the past 12 months using a screening questionnaire (19). Non-asthmatics will have had no asthma diagnosis and no current/previous asthma symptoms (Table 1).

**Table 1:**
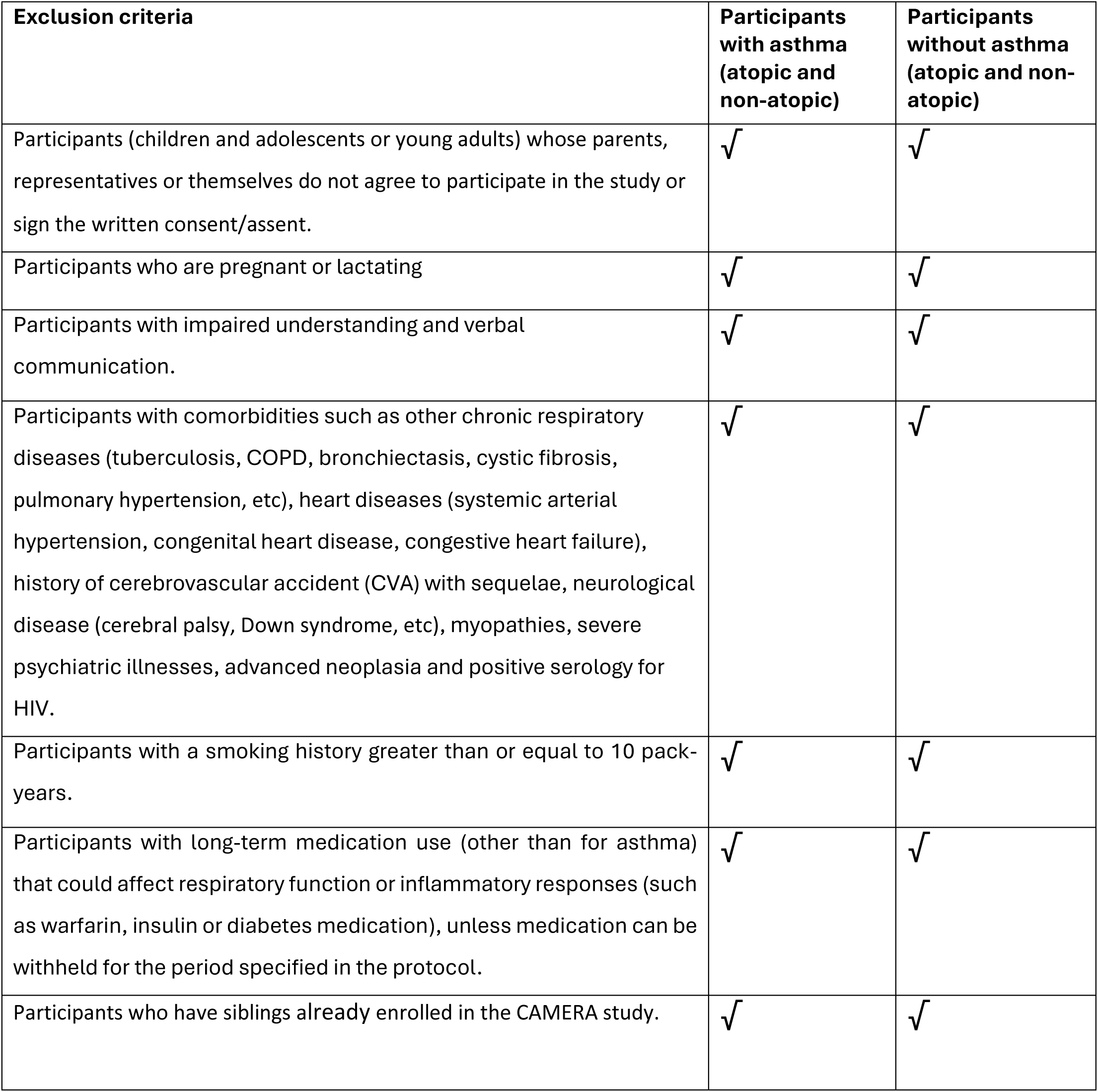
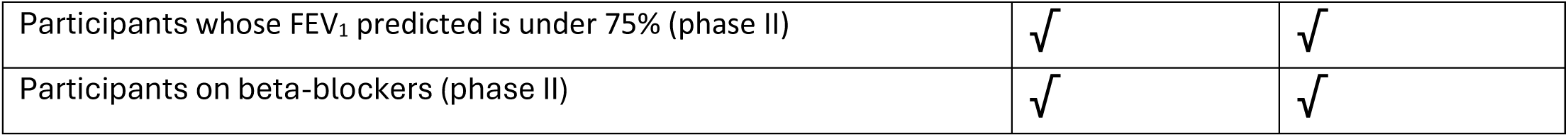
General exclusion criteria for asthmatic and non-asthmatic participants.

| **Exclusion criteria** | **Participants with asthma (atopic and non-atopic)** | **Participants without asthma (atopic and non-atopic)** |
| --- | --- | --- |
| Participants (children and adolescents or young adults) whose parents, representatives or themselves do not agree to participate in the study or sign the written consent/assent. | √ | √ |
| Participants who are pregnant or lactating | √ | √ |
| Participants with impaired understanding and verbal communication. | √ | √ |
| Participants with comorbidities such as other chronic respiratory diseases (tuberculosis, COPD, bronchiectasis, cystic fibrosis, pulmonary hypertension, etc), heart diseases (systemic arterial hypertension, congenital heart disease, congestive heart failure), history of cerebrovascular accident (CVA) with sequelae, neurological disease (cerebral palsy, Down syndrome, etc), myopathies, severe psychiatric illnesses, advanced neoplasia and positive serology for HIV. | √ | √ |
| Participants with a smoking history greater than or equal to 10 pack-years. | √ | √ |
| Participants with long-term medication use (other than for asthma) that could affect respiratory function or inflammatory responses (such as warfarin, insulin or diabetes medication), unless medication can be withheld for the period specified in the protocol. | √ | √ |
| Participants who have siblings already enrolled in the CAMERA study. | √ | √ |
| Participants whose FEV<sub>1</sub> predicted is under 75% (phase II) | √ | √ |
| Participants on beta-blockers (phase II) | √ | √ |

#### Data collection

Data will be collected using standardised instruments and clinical tests using validated protocols. Questionnaires will assess respiratory health, asthma control (20) and perceived stress (21). Blood, hair and nasal samples will be collected. Respiratory assessments will include lung function testing, capsaicin challenge, dry air challenge, and cold air challenge.

#### Questionnaires

The asthma and triggers questionnaire (adapted from the ISAAC phase II questionnaire (19)) will collect data about respiratory health (asthma history, symptoms and triggers) and medication use in the year before recruitment, active smoking and vaping, and general health. The Asthma Control Questionnaire will collect data on current asthma symptoms and management (20) from asthma cases only. A questionnaire based on the standard Cohen questionnaire (21) will be administered on the day of hair sample collection to evaluate participants’ stress levels in the last four weeks, the occurrence of potential stressful events and whether they have induced or worsened respiratory symptoms.

#### Skin Prick Test (SPT)

The SPT will follow a standardised protocol (22). Histamine and saline will be used as positive and negative controls, respectively. A positive reaction will be defined as a mean wheal size of ≥3mm (after subtracting the negative control wheal size) fifteen minutes after the test. A positive reaction to at least one allergen from the panel will be considered a positive SPT. The panel includes house dust mite, cat/dog dander, mixed tree pollen and centre-specific allergens (Immunotech USA and Inmunotek Europe) (Table S1).

#### Lung Function Test (LFT)

LFT will be conducted according to American Thoracic Society (ATS) criteria (23). Spirometry will be performed using portable ultrasonic devices (e.g. EasyOne ndd Medical Technologies, Zurich, Switzerland). All spirometers will undergo regular tests by the suppliers. Participants will maintain a sitting position during the test, and the best of three reproducible forced expiratory manoeuvres will be used to obtain the following lung function parameters: Forced Vital Capacity (FVC), Forced Expiratory Volume in 1 second (FEV_1_), FEV/FVC ratio, Maximal Mid-Expiratory Flow (MMEF) and Peak Expiratory Flow (PEF).

#### Nasal lavage

Nasal lavage will be conducted using a standardised protocol (24). Mucosal Atomisation Devices (MAD) will be used to deliver sterile saline or phosphate buffered saline (PBS) solution (2 ml) into each nostril, and the sample will be collected using a conical tube/funnel once the participant blows strongly through the nostril into the funnel; this will be repeated for the other nostril. Cell suspensions will be counted, centrifuged, and stained (Diff-Quik; Dade Behring, Deerfield, Illinois, USA). The laboratory in Brazil will complete the inflammatory cell counts for all centres.

#### Blood sampling

10 mL of venous blood will be collected using Vacutainer tubes (Sodium heparin and serum separator tube, Becton Dickinson) for eosinophil counts, whole blood culture for IFN-γ, and total and allergen-specific IgE (asIgE) measurement. Samples will undergo processing within 4-6 hours of collection.

#### IFN-γ

Heparinised whole blood will be diluted at a 1:4 ratio in Roswell Park Memorial Institute (RPMI) medium containing 10 mM glutamine and 100 ug/mL gentamicin in 0.5 ml tissue culture wells and maintained at 37^0^C in a humidified 5% CO_2_ incubator for five days. Culture conditions will include unstimulated (medium alone), pokeweed mitogen (Sigma/Merck) (2.5µg/ml), and *Dermatophagoides pteronyssinus* antigen (LoTox Der p 1, 20µg/mL, catalog number: LTN-DP1-1, Inbio Charlottesville, USA). Supernatants will be collected into 1.5 mL tubes and stored at -70°C until analysis. Supernatants will be analysed using an IFN-γ enzyme-linked immunosorbent assay (ELISA) following the manufacturer’s instructions (R&D Systems/ BD OptEIA™ Phamingen San Diego, USA).

#### Hair cortisol

Hair samples (approximately 50-350mg or 1cm long) will be collected from the posterior vertex region of the scalp, cut close to the scalp surface, stored in specimen paper within resealable envelopes, and maintained at room temperature in dark conditions (25). Cumulative cortisol concentration, a stress biomarker (26), will be measured from ≥10 mg of hair cut from the scalp end per sample. The sample will be cut into pieces and incubated overnight in 1ml methanol at 52°C. The resulting supernatant will be air-dried in a refrigerator and reconstituted in PBS based on sample weight. Cortisol levels in the hair supernatant will be quantified using an ELISA cortisol kit (*Biomatik* Cortisol (Cor) ELISA kit, cat# EKU03476, Wilmington, USA).

#### Dry air challenge

Bronchial provocation using dry mixed air (21% O_2_, 74% N_2,_ with 5% CO_2_ to prevent dizziness) will follow a well-defined protocol (27). Spirometry will be conducted to measure baseline FEV_1_, and the challenge will only be conducted if FEV_1_ ≥75% predicted value. Dry mixed air will be continuously provided through two 20L breathing bags, with participants inhaling for six minutes (via mouthpiece with nose clipped). During this time, participants will increase and sustain their ventilation rate at 60% of the maximum voluntary ventilation (MVV; or FEV_1_ × 35), with an acceptable ventilation range of 16-25 times baseline FEV_1_ (28,29). The minute ventilation rate will be monitored and guided using a ventilation meter (TAC-7715, Equilibrated Bio Systems, Inc., Melville, New York, USA); verbal guidance will be provided throughout the test. For safety, a fingertip pulse oximeter will be used throughout to ensure oxygen saturation levels (saO_2_) remain above 85%. After the challenge, participants will undergo spirometry at 5, 10, 15 and 30 minutes. A ≥10% fall in FEV_1_ post-challenge will be considered a positive eucapnic voluntary hyperpnea (EVH) test (30). Salbutamol (200 mcg) will be administered if there is a FEV_1_ drop ≥ 15%. Participants will be monitored closely post-challenge for any adverse reactions using centre-specific safety assessment procedures.

#### Capsaicin challenge

Capsaicin challenge using inhaled capsaicin solution (Stockport Pharmaceuticals, UK) will be conducted based on European Respiratory Society (ERS) guidelines (31). Solutions will be prepared daily following a standardised capsaicin dilution protocol, with doubling concentrations (0.98 – 500 μM) and a negative (saline) control (32). With participants in a sitting position, a compressed air-driven jet nebuliser controlled by a dosimeter (ProvoX, Ganshorn, Germany) will be used to administer single doses of aerosolised capsaicin solution via a mouthpiece. After dose administration, the number of coughs during the subsequent 30 seconds will be recorded. After this period, urge to cough (using a Borg scale) and instances of throat clearing (TC) will also be recorded. This will then be repeated with the increasing capsaicin dosages, with a one-minute delay before administering the next concentration. The lowest concentration that elicits two and five coughs, C2 and C5, respectively, will be recorded (33). For safety, spirometry will be conducted at baseline, mid-point (for participants with dyspnoea) and post-challenge. Salbutamol (200 mcg) will be administered if FEV_1_ drops below 75% of predicted. Participants will be monitored closely post-challenge for any adverse reactions using centre-specific safety assessment procedures.

#### Cold air challenge

Bronchial provocation will be assessed in NZ and Uganda using cold, dry mixed air according to a well-defined existing protocol (27,34). Following baseline spirometry, participants will inhale cold (- 20°c), dry air (21% O_2_, 74% N_2_, with 5% CO_2_ to prevent dizziness) for 3 minutes at rest. They will then be asked to increase ventilation rate (hyperventilate) at a target ventilation rate of 60% of MVV, with an acceptable range of ventilation of 16-25 times baseline FEV_1_ during the challenge (35). The minute ventilation rate and safety will be monitored and guided as described for the dry air challenge. Participants will undergo spirometry at 5, 10, 15 and 30 minutes post challenge and a positive challenge response will be defined as a 10-15% decrease in FEV1.

### Data Management and Analysis

Electronic data will be collected using Kobo Toolbox or REDCap (Research Electronic Data Capture) databases. For NZ, data will be backed up using Microsoft Access database. Centre-specific quality control measures include mandatory completion of electronic fields, medical record review and completion verification within 48 hours of each visit, and internal data audits before data transfer. Pseudonymised data, identifiable by participants’ identification number and date of birth (age in years), will be transferred securely to the London School of Hygiene and Tropical Medicine (LSHTM) for centralised processing and review. Data cleaning and quality checks will be conducted regularly by LSHTM researchers according to a detailed data transfer guide and study manual.

Data will be analysed using STATA 18 (36) and R v 4.3.3 (37). Descriptive analyses will be conducted to describe AA/NAA and atopic/non-atopic non-asthmatics using demographic data, respiratory health, asthma characteristics (triggers, control, treatment), active smoking, baseline/post-challenge predicted lung function parameters, SPT/asIgE data, and blood eosinophils. Primary analyses will compare continuous outcomes from test data collected in the four study groups within each centre. Statistical analysis will include t-tests to compare geometric means of capsaicin cough concentrations, inflammatory markers and IFN-γ levels; generation of standard hair cortisol concentration curves; comparison of FEV_1_ reductions from baseline values for dry and cold air challenges, across study groups and centres; evaluation of capsaicin cough concentrations by country, ethnicity, and asthma medication use. We will also assess differences in FEV_1_ responses between dry and cold air challenges in NZ and Uganda.

Multivariable linear regression adjusting for study centre and potential confounders will be used to examine associations between mechanistic outcomes and atopy/non-atopy. We will test if associations differ by study centre and stratify if indicated. Any between-centre differences will be explored, accounting for variations in geographic settings, environmental exposures, socioeconomic factors and asthma prevalence.

### Study size and power

Based on previous estimates (17), the CAMERA study will have 80% power to detect a 0.65 standard deviation difference between groups for continuous outcomes, with 40 participants per group (AA, NAA, atopic non-asthmatics, non-atopic non-asthmatics) per centre. Pooling findings across centres will further increase the power to detect smaller differences. For binary outcomes, assuming a 25% outcome in one group and 50% in another, there will be 82% power to detect a 50% difference in proportions between different groups, with 160 participants in each of the four groups of interest overall.

## Ethical considerations

Ethical approval was obtained from LSHTM (ref: 26308 - 1) and all participating centres prior to commencing the study as follows: the Brazilian National Research Ethics Council (number 59164922.6.0000.5577), Hospital San Francisco de Quito (Unique Approval Ethics Committee Code CEISH-HGSF-2022-023), NZ Health and Disability Ethics Committees (ref: 2022 EXP 12986) and Uganda National Council for Science and Technology (ref: 2022 EXP 12986).

## Discussion

In light of the poorly understood and understudied mechanisms of NAA and degree of overlap with non-eosinophilic asthma in LMICs and HICs, the findings of this multi-centre study will inform new and phenotype-specific prevention and treatment approaches. Identifying different mechanisms (i.e. physiological, inflammatory or neural) could help inform policies around non-pharmacological approaches, such as physical exercise/weight loss (38), improving indoor air quality (38), smoking cessation, and stress management techniques (3), to tackling NAA in different settings. Understanding neural mechanisms could help guide the development of novel pharmacological interventions targeting non-eosinophilic inflammatory mechanisms in asthma. By leveraging multidimensional data, including inflammatory markers, lung function, and neural sensitivity tests, we aim to uncover previously unrecognised mechanisms driving NAA in diverse populations.

Given that this is a multi-country study, we have carefully standardised methods and protocols for each procedure for consistency, and have conducted extensive procedure training, quality control, and training on equipment use and assembly and specific clinical tests and challenges. This study promotes equitable, ethical and mutually beneficial partnerships ensuring that the main outcomes align with local public health needs and priorities. The data management and analysis process aims to leverage local resources equitably while respecting intellectual property rights and adhering to local regulations.

Ultimately, we will use these standardised protocols to compare geographically diverse populations and will incorporate challenge tests beyond those typically included in epidemiological studies to focus on the poorly understood but very important NAA phenotype. The CAMERA protocol incorporates more detailed mechanistic investigations compared to previous studies, such as the WASP study (16). This study will provide novel insights into the mechanisms of NAA across HICs and LMICs and potentially help address the disproportionate burden of NAA across different geographical and income settings.

To our knowledge, this observational study will be the first to provide an in-depth evaluation of mechanisms underlying NAA in children and young adults globally. We are presenting this protocol to allow reproducibility of the methods and procedures, and to enable other researchers to conduct studies within HICs or LMICs using a standardised protocol.

## Supporting information

Supplemental Table 1 and Figure 1, and will be used for the link to the file on the preprint site.

## Data Availability

As this study is currently ongoing, no data are available at this time.

## Acknowledgements

We are grateful to the returning WASP and new CAMERA participants, researchers and lab technicians for conducting clinical tests and sample processing, LSHTM researchers for data management and analysis and the hospitals and institutions at ProAR, Massey University, MRC Uganda and Manabí, Ecuador, for allowing CAMERA researchers to use their facilities during phase II of the study.

## Support statement

The CAMERA project was supported by the European Research Council under ERC grants 668954 and 101020088.

The CAMERA Study group:

<u>United Kingdom, London:</u> Neil Pearce, Lucy Pembrey, Mary Njoroge, Steven Robertson, Charlotte E Rutter, Donna Davoren, David Rebellon Sanchez

<u>Brazil:</u> Alvaro Cruz, Camila Figueiredo, Mauricio Barreto, Cinthia Vila Nova Santana, Gabriela Pimentel Pinheiro, Gilvaneide Lima, Leticia Marques, Auxiliadora Damiane Costa, Karla Nicole, Candace Andrade, Bianca Sampaio Dotto Fiuza, Luciano Gama da Silva Gomes,, Louise Correia de Lima, Mel Matos de Carvalho Espinheira, Manoela Souza Trindade Fontes, Carolina Barbosa Souza Santos, Emile Ivana Fernandes Santos Costa, Rayane Anselmo dos Santos Ribeiro, Thalia Raney Alves Pereira Santos.

<u>Ecuador:</u> Philip Cooper, Natalia Romero-Sandoval, Santiago Mena, Lucia Alonzo, Genesis Palma, Maria Fernanda Arias, Jessica Alchundia, Karla Solorzano, Martha Chico.

<u>New Zealand:</u> Jeroen Douwes, Collin Brooks, Hajar Ali, Jeroen Burmanje, Stephanie Hobbs

<u>Uganda:</u> Harriet Mpairwe, Irene Nambuya, Pius Tumwesige, Milly Namutebi, Marble Nnaluwooza, Mike Mukasa

