## Supplemental Table 1 and Figure 1, and will be used for the link to the file on the preprint site. for "Causes And MEchanisms foR non-atopic Asthma in Children (CAMERA) Study: rationale and protocol"

**Supplementary figures and tables**

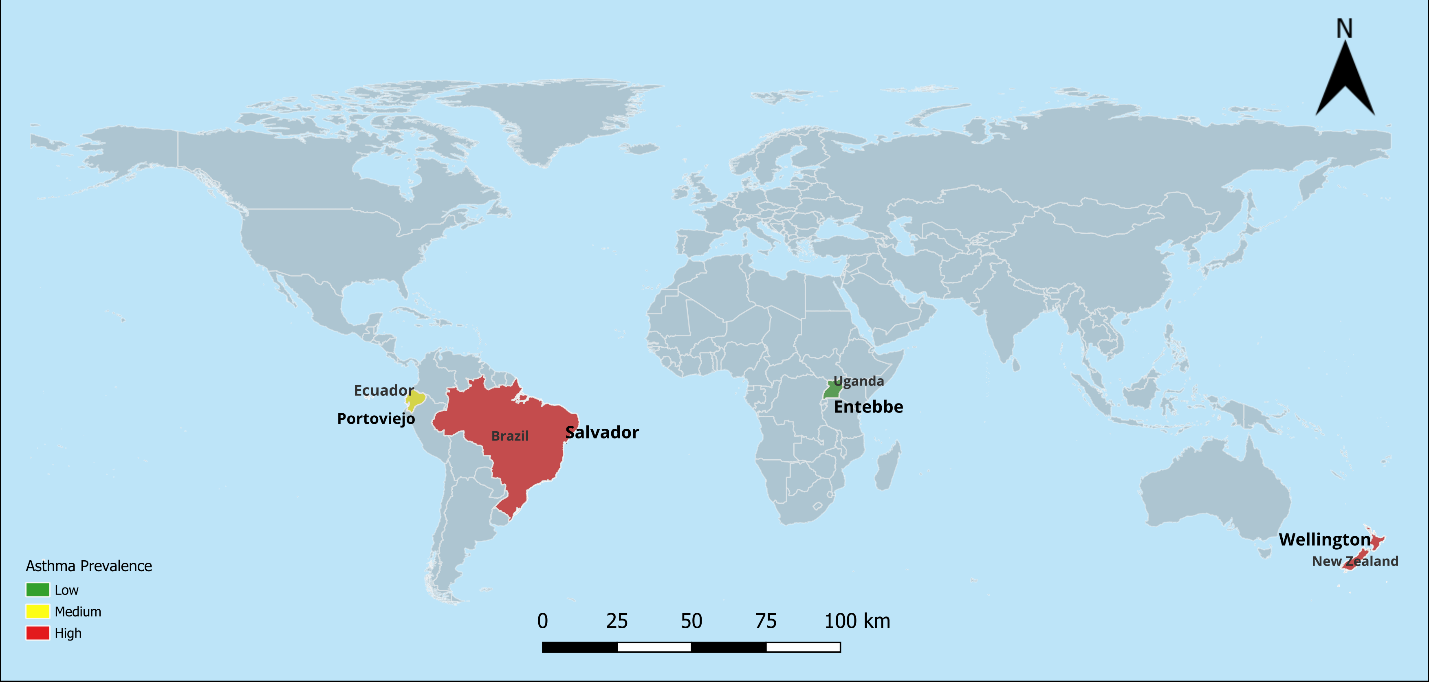

Figure S1: Map showing the location of CAMERA study centres by country and asthma prevalence

^*Brazil, Ecuador, and Uganda are LMICs whereas New Zealand is a HIC^

Table S1:Skin prick test allergens to be used in each study centre

| **Allergen list** | **Brazil** | **New Zealand** | **Ecuador** | **Uganda** |
| --- | --- | --- | --- | --- |
| (1) Negative control (saline)^*^ | √ | √ | √ | √ |
| (2) Grass mix |  | √ |  |  |
| (3) House dust mite (*Dermatophagoides pteronyssinus*)^*^ | √ | √ | √ | √ |
| (4) Cat^*^ | √ | √ | √ | √ |
| (5) Dog^*^ | √ | √ | √ | √ |
| (6) Mixed tree pollen^*^ | √ | √ | √ | √ |
| (7) House dust mite (*Dermatophagoides farinae*) | √ |  | √ |  |
| (8) House dust mite (*Dermatophagoides* mix) |  |  |  | √ |
| (9) *Cladosporium herbarum* (mould) |  |  | √ |  |
| (10) *Aspergillus fumigatus* (mould) |  |  | √ |  |
| (11) Weed mix |  |  |  | √ |
| (12) *Alternaria alternata* (mould mix) |  |  |  | √ |
| (13) Feather mix |  |  |  | √ |
| (14) *Blomia tropicalis* | √ |  | √ | √ |
| (15) *Blatela germanica* (cockroach) | √ |  | √ |  |
| (16) *Periplaneta Americana* (cockroach) | √ |  | √ | √ |
| (17) Positive control (histamine)^*^ | √ | √ | √ | √ |

^*Core allergens to be tested^

**C**auses **A**nd **ME**chanisms fo**R** non-atopic **A**sthma in children (CAMERA)

Participant Questionnaire

### PLEASE READ THIS CAREFULLY BEFORE BEGINNING THE QUESTIONNAIRE

Thank you for agreeing to complete this questionnaire. We are very grateful for your ongoing contribution, and this information is extremely valuable to our research. The questions should be answered by the participant but you may check some details with parents/guardians if necessary.

- Most questions require you to TICK 🗹 your answer in a box. If you make a mistake put a cross in the box and tick the correct answer. Only tick one option unless otherwise instructed.
- Examples of how to mark the questionnaire.

To answer “yes”	🗹 Yes	🞎 No

To answer “no”	🞎 Yes	🗹 No

If you have any queries or problems with the completion of this questionnaire, please contact us for assistance on: Email: [local organiser email address]	**or**

Telephone: [local organiser phone number]

### CONSENT

#### Has the participant consented to participate in this study? If yes, date consented

| d | d | m | m | y | y | y | y |
| --- | --- | --- | --- | --- | --- | --- | --- |

Yes 🞎	No 🞎

### DEMOGRAPHIC INFORMATION OF CHILD/ADOLESCENT PARTICIPATING IN THIS STUDY

CAMERA Study ID:

Name:

Date questionnaire completed: Full date of birth:

| d | d | m | m | y | y | y | y |
| --- | --- | --- | --- | --- | --- | --- | --- |
| d | d | m | m | y | y | y | y |

| **1** | **5** | m | m | y | y | y | y |
| --- | --- | --- | --- | --- | --- | --- | --- |

Estimated using 15th of month:

(Use this box where only month and year are known)

What was your sex at birth?

- Male
- Female
- Other (specify):

Is the gender that you identify with the same as your sex registered at birth?

- Yes
- No	If No, please specify gender identity:

Height (cm):

Weight (kg):

### RESPIRATORY HEALTH AND ALLERGIES

1. Have you ever had any of the following at any time in the past? Wheeze (a whistling sound from the chest)

| Yes 🞎 | No 🞎 |
| --- | --- |
| Yes 🞎 | No 🞎 |
| Yes 🞎 | No 🞎 |
| Yes 🞎 | No 🞎 |
| Yes 🞎 | No 🞎 |
| Yes 🞎 | No 🞎 |

Cough

Shortness of breath Chest tightness Rhinitis

Eczema

2. Have you had wheezing or whistling in your chest at any time in the past 12 months?
   - Yes

#### No. IF YOU HAVE ANSWERED ‘*NO*’, PLEASE GO TO QUESTION 9.

3. How many attacks of wheezing have you had in the past 12 months?
   - None
   - 1-3 times
   - 4-12 times
   - More than 12 times
4. In the past 12 months, how often, on average, has your sleep been disturbed due to wheezing?
   - Never woken with wheezing
   - Less than one night per week
   - One or more nights per week
5. In the past 12 months, has wheezing ever been severe enough to limit your speech to only one or two words at a time between breaths?
   - Yes
   - No
6. In the past 12 months, has your chest sounded wheezy during or after exercise?
   - Yes
   - No
7. Have you been at all breathless when the wheezing noise was present?
   - Yes
   - No
8. In the past 12 months, have you had a wheezing or whistling in the chest when you did not have a cold?
   - Yes
   - No
9. In the past 12 months, have you had a dry cough at night, apart from a cough associated with a cold or chest infection?
   - Yes
   - No
10. Do you often cough at night when you don’t have a cold or the flu?
    - Yes
    - No
11. How often, on average, has your sleep been disturbed due to coughing?
    - Never disturbed due to coughing
    - Less than one night per week
    - One or more nights per week

#### These questions are about problems which occur when you DO NOT have a cold or the flu.

12. In the past 12 months have you had a problem with sneezing or a runny or blocked nose when you **DID NOT** have a cold or the flu?
    - Yes

#### No IF YOU HAVE ANSWERED ‘*NO*’, PLEASE GO TO QUESTION 16.

13. In the past 12 months, has this nose problem been accompanied by itchy-watery eyes?
    - Yes
    - No
14. In which of the past 12 months did this nose problem occur?

| January | Yes 🞎 | No 🞎 | July | Yes 🞎 | No 🞎 |
| --- | --- | --- | --- | --- | --- |
| February | Yes 🞎 | No 🞎 | August | Yes 🞎 | No 🞎 |
| March | Yes 🞎 | No 🞎 | September | Yes 🞎 | No 🞎 |
| April | Yes 🞎 | No 🞎 | October | Yes 🞎 | No 🞎 |
| May | Yes 🞎 | No 🞎 | November | Yes 🞎 | No 🞎 |

June

Yes 🞎	No 🞎

December

Yes 🞎	No 🞎

15. In the past 12 months, how much did this nose problem interfere with your daily activities?
    - Not at all
    - A little
    - A moderate amount
    - A lot
16. In the past 12 months, have you had an itchy rash which was coming and going for at least six months?
    - Yes

#### No IF YOU HAVE ANSWERED ‘*NO*’, PLEASE GO TO QUESTION 20.

17. Has this itchy rash at any time affected any of the following places: the folds of the elbows, behind the knees, in front of the ankles, under the buttocks, or around the neck, ears or eyes?
    - Yes
    - No
18. Has this rash cleared completely at any time during the past 12 months?
    - Yes
    - No
19. In the past 12 months, how often, on average, have you been kept awake at night by this itchy rash?
    - Never in the past 12 months
    - Less than one night per week
    - One or more nights per week
20. Have you been woken by an attack of shortness of breath at any time in the past 12 months?
    - Yes
    - No
21. Have you woken up with a feeling of tightness in your chest at any time in the past 12 months?
    - Yes
    - No
22. In the past 12 months, how often have you been unable to attend school or prevented from doing normal activities because of respiratory symptoms i.e. cough, phlegm, wheezing/whistling or shortness of breath?
    - Never
    - 1-7 days
    - 8-30 days
    - At least 31 days
    - Don’t know
23. Have you ever had asthma?
    - Yes

#### No IF YOU HAVE ANSWERED ‘*NO*’, PLEASE GO TO QUESTION 31.

24. Was the diagnosis confirmed by a doctor?
    - Yes	If ***‘YES’*** at what age was the diagnosis made*?*	Years
    - No
25. How old were you when you had the first attack of asthma?	Years
26. How old were you when you had the last attack of asthma?	Years
27. Have you had an attack of asthma in the past 12 months?
    - Yes
    - No
28. Have you ever visited a hospital A & E (Accident and Emergency) Department or after hours General Practitioner /medical centre because of your breathing?
    - Yes
    - No
29. Have you ever spent a night in hospital because of breathing problems?
    - Yes
    - No
30. How many nights have you spent in hospital in the past 12 months?	Nights
31. In the past 12 months, have you taken any medication, pills, inhalers, or other medications for asthma/wheeze/other?

🞎 Yes	🞎 No	**If *NO*, please go to question 34.**

If ***YES*,** Please specify:

Asthma Wheeze Other

🞎 Yes	🞎 No

🞎 Yes	🞎 No

🞎 Yes	🞎 No

If YES specify other:

32. Select or list the medication(s) that you have taken in the past 12 months, and how often.

When wheezy	Regularly

Reliever (blue inhaler)

🞎 Yes	🞎 No If yes:	🞎	🞎

Preventer (brown/beige/white/red/orange inhalers) 🞎 Yes	🞎 No If yes:	🞎	🞎

Antihistamines

Nasal spray **(If yes, please complete question 33)**

🞎 Yes	🞎 No. If yes:	🞎	🞎

🞎 Yes	🞎 No If yes:	🞎	🞎

| Oral corticosteroids (e.g. Prednisolone) | - Yes | - No | If yes: | 🞎 🞎 |
| --- | --- | --- | --- | --- |
| Others (*please list*) | - Yes | - No |  |  |
| 1. |  |  | If yes: | 🞎 🞎 |
| 2. |  |  | If yes: | 🞎 🞎 |
| 3. |  |  | If yes: | 🞎 🞎 |
| 4.  5. |  |  | If yes:  If yes: | 🞎 🞎  🞎 🞎 |

33. If you have used a nose spray, what are the names of nose sprays that you have used?

1.

2.

3.

4.

5.

6.

7.

34. Have you ever received treatment with any of the following to treat parasites or protect against Coronavirus (SARS-CoV-2)?
a) Ivermectin (local commercial name for drug)	Yes 🞎	No 🞎	Don’t know/not sure 🞎

If yes, have you received this treatment to treat parasites or protect against Coronavirus

(SARS-CoV-2) in the past 12 months?

b) Albendazole (local commercial name for drug)

Yes 🞎	No 🞎	Don’t know/not sure 🞎

Yes 🞎	No 🞎	Don’t know/not sure 🞎

If yes, have you received this treatment to treat parasites or protect against Coronavirus

(SARS-CoV-2) in the past 12 months?

c) Praziquantel (local commercial name for drug)

Yes 🞎	No 🞎	Don’t know/not sure 🞎

Yes 🞎	No 🞎	Don’t know/not sure 🞎

If yes, have you received this treatment to treat parasites or protect against Coronavirus

(SARS-CoV-2) in the past 12 months?

d) Other

If yes, please specify:

Yes 🞎	No 🞎	Don’t know/not sure 🞎

Yes 🞎	No 🞎	Don’t know/not sure 🞎

If yes, have you received this treatment to treat parasites or protect against Coronavirus

(SARS-CoV-2) in the past 12 months?

Yes 🞎	No 🞎	Don’t know/not sure 🞎

### ACTIVE SMOKING

35. Have you ever smoked cigarettes, cigars or other (excluding e-cigarettes/vapes)?
    - Yes
    - No	If *No*, please go to question 40.
36. Have you smoked more than 5 packs of cigarettes in total in your whole life?
    - Yes
    - No
37. At what age did you start smoking?	Years
38. Do you still smoke?
    - Yes
    - No
39. What did you or do you smoke and how many per day?

If Yes, number smoked per day

Cigarettes Cigars Other:

🞎 Yes	🞎 No

🞎 Yes	🞎 No

🞎 Yes	🞎 No

(If yes, specify other)

[per day]

[per day]

[per day]

40. Have you ever used e-cigarettes, e.g. vapes?
    - Yes	If yes, how long have you used e-cigarettes/vaping devices for?
    - No (Go to Q42)
    - <1 month
    - 1-3 months
    - 4-6 months
    - 7 months to 1 year
    - 1-2 years
    - >2 years
41. How often do you use electronic cigarettes/vaping devices?
    - At least once a day
    - At least once a week
    - At least once a month
    - Less than once a month

### ASTHMA TRIGGERS AND GENERAL HEALTH

42. What triggers seem to make your asthma symptoms worse?

| Exercise | - Yes | - No | - Not sure |
| --- | --- | --- | --- |
| Chest infection | - Yes | - No | - Not sure |
| Air pollution/irritants | - Yes | - No | - Not sure |
| Cold air | - Yes | - No | - Not sure |
| Dust | - Yes | - No | - Not sure |
| Laughter/Psychological stress | - Yes | - No | - Not sure |
| Pollen or pets | - Yes | - No | - Not sure |
| Do not know | - Yes | - No | - Not sure |
| Other | - Yes | - No | - Not sure |
| If other, please specify: |  |  |  |

43. In general, would you say your health is?
    - Excellent
    - Very good
    - Good
    - Fair
    - Poor
44. Compared to one year ago, how would you rate your health in general now?
    - Much better now than 1 year ago
    - Somewhat better now than 1 year ago
    - About the same as 1 year ago
    - Somewhat worse now than 1 year ago
    - Much worse now than 1 year ago

Researcher name:

Researcher signature:

**THANK YOU VERY MUCH FOR COMPLETING THE QUESTIONNAIRE!**

**Asthma Control Questionnaire**

CAMERA Study ID:	Date questionnaire completed:

| d | d | m | m | y | y | y | y |
| --- | --- | --- | --- | --- | --- | --- | --- |

| d | d | m | m | y | y | y | y |
| --- | --- | --- | --- | --- | --- | --- | --- |

Full date of birth:

Estimated using 15th of month:

| **1** | **5** | m | m | y | y | y | y |
| --- | --- | --- | --- | --- | --- | --- | --- |

(Use this box where only month and year are known)

1. On average, during the past week, how often were you woken up by your asthma during the night?
2. On average, during the past week, how bad were your asthma symptoms when you woke up in the morning?
3. In general, during the past week, how limited were you in your activities because of your asthma?
   - Never
   - Hardly ever
   - A few times
   - Several times
   - Many times
   - A great many times
   - Unable to sleep because of asthma
   - No symptoms
   - Very mild symptoms
   - Mild symptoms
   - Moderate symptoms
   - Quite severe symptoms
   - Severe symptoms
   - Very severe symptoms
   - Not limited at all
   - Very slightly limited
   - Slightly limited
   - Moderately limited
   - Very limited
   - Extremely limited
   - Totally limited
4. In general, during the past week, how much shortness of breath did you experience because of your asthma?
5. In general, during the past week, how much of the time did you wheeze?
6. On average, during the past week, how many puffs/inhalations of short-acting bronchodilator have you used each day?
   - None
   - A very little
   - A little
   - A moderate amount
   - Quite a lot
   - A great deal
   - A very great deal
   - Never
   - Hardly any of the time
   - A little of the time
   - A moderate amount of the time
   - A lot of the time
   - Most of the time
   - All the time
   - None
   - 1-2 puffs/inhalations most days
   - 3-4 puffs/inhalations most days
   - 5-8 puffs/inhalations most days
   - 9-12 puffs/inhalations most days
   - 13-16 puffs/inhalations most days
   - More than 16 puffs/inhalations most days

Researcher name:

Researcher signature:

**Asthma stress questionnaire**

**C**auses **A**nd **ME**chanisms fo**R** non-atopic **A**sthma in children (CAMERA)

**Perceived Stress Questionnaire**

**PLEASE READ THIS CAREFULLY BEFORE BEGINNING THE QUESTIONNAIRE**

Thank you for agreeing to complete this questionnaire. We are very grateful for your ongoing contribution, and this information is extremely valuable to our research. The questions should be answered by the participant but you may check some details with parents/guardians if necessary.

CAMERA Study ID:	Full date of birth:

| d | d | m | m | y | y | y | y |
| --- | --- | --- | --- | --- | --- | --- | --- |

| **1** | **5** | m | m | y | y | y | y |
| --- | --- | --- | --- | --- | --- | --- | --- |

Estimated using 15th of month:

(Use this box where only month and year are known)

Date Perceived Stress Questionnaire completed:

| d | d | m | m | y | y | y | y |
| --- | --- | --- | --- | --- | --- | --- | --- |

### THE FOLLOWING QUESTIONS ARE SENSITIVE AND PERSONAL.

The following questions aim to assess whether stressful events have occurred and whether they may have contributed to the development or worsening of respiratory symptoms. This is highly relevant since stressful events have previously been shown to be, at least in part, responsible for asthma attacks in some people. Given the sensitive nature we understand that some participants may wish to decline answering these questions. If this is the case, then feel free to not complete all, or part of this section.

### PERCEIVED STRESS

The questions in this section will ask you about your feelings and thoughts ***during the last month.***

1. In the last month, how often have you been upset because of something that happened unexpectedly?

- Never
- Almost never
- Sometimes
- Very often

2. In the last month, how often have you felt that you were unable to control the important things in your life?

- Never
- Almost never
- Sometimes
- Very often

3. In the last month, how often have you felt nervous and "stressed"?

- Never
- Almost never
- Sometimes
- Very often

4. In the last month, how often have you dealt successfully with irritating life hassles?

- Never
- Almost never
- Sometimes
- Very often

5. In the last month, how often have you felt that you were effectively coping with important changes that were occurring in your life?

- Never
- Almost never
- Sometimes
- Very often

6. In the last month, how often have you felt confident about your ability to handle your personal problems?

- Never
- Almost never
- Sometimes
- Very Often

7. In the last month, how often have you felt that things were going your way?

- Never
- Almost never
- Sometimes
- Very often

8. In the last month, how often have you found that you could not cope with all the things that you had to do?

- Never
- Almost never
- Sometimes
- Very often

9. In the last month, how often have you been able to control irritations in your life?

- Never
- Almost never
- Sometimes
- Very often

10. In the last month, how often have you felt that you were on top of things?

- Never
- Almost never
- Sometimes
- Very often

11. In the last month, how often have you been angered because of things that happened that were outside of your control?

- Never
- Almost never
- Sometimes
- Very often

12. In the last month, how often have you found yourself thinking about things that you have to accomplish?

- Never
- Almost never
- Sometimes
- Very often

13. In the last month, how often have you been able to control the way you spend your time?

- Never
- Almost never
- Sometimes
- Very often

14. In the last month, how often have you felt difficulties were piling up so high that you could not overcome them?

- Never
- Almost never
- Sometimes
- Very often

Researcher name:

Researcher signature:

### THANK YOU VERY MUCH FOR COMPLETING THE QUESTIONNAIRE!
